# Natural Language Processing to Identify Racial and Ethnic Disparities in Aortic Stenosis

**DOI:** 10.1101/2023.12.15.23300011

**Authors:** Dhruva Biswas, Jack Wu, Apurva Bharucha, Natalie Fairhurst, George Kaye, Kate Jones, Freya Parker Copeland, Bethan O’Donnell, Daniel Kyle, Tom Searle, Nilesh Pareek, Rafal Dworakowski, Alexandros Papachristidis, Narbeh Melikian, Olaf Wendler, Ranjit Deshpande, Max Baghai, James Galloway, James T Teo, Richard Dobson, Jonathan Byrne, Philip MacCarthy, Ajay M Shah, Mehdi Eskanderi, Kevin O’Gallagher

**Affiliations:** School of Cardiovascular and Metabolic Medicine & Sciences, King’s College London, UK; Cardiovascular Department,King’s College Hospital NHS Foundation Trust, London, UK; South London Office of Specialised Services, South London Cardiovascular Network; Cleveland Clinic, London, UK; Centre for Rheumatic Disease, King’s College London; Department of Rheumatology, King’s College Hospital NHS Foundation Trust, London, UK; Institute of Psychiatry, Psychology and Neurosciences, King’s College London, UK; Department of Neurology, King’s College Hospital NHS Foundation Trust.

## Abstract

**IMPORTANCE:** This study uses artificial intelligence (AI) technologies to augment quality measurement and improvement in the setting of aortic stenosis (AS). We characterise racial and ethnic disparities in the diagnosis, management, and outcome of AS within a universal healthcare system.

**OBJECTIVE:** To use natural language processing (NLP) AI methods applied to the electronic health records (EHR) to identify racial and ethnic disparities in AS while correcting for the effects of socioeconomic deprivation.

**DESIGN:** Retrospective cohort study.

**SETTING:** King’s College Hospital NHS Foundation Trust, a multi-site tertiary care hospital in London, UK

**PARTICIPANTS:** Adult patients with a diagnosis of AS between 2010-2020.

**MAIN OUTCOMES AND MEASURES:** Key outcomes were all-cause mortality, frequency of AS intervention (TAVI or surgical aortic valve replacement [AVR]) and the time from diagnosis of severe AS to intervention. All analyses were adjusted for age, sex and socioeconomic deprivation.

**RESULTS:** 5859 patients with AS were identified, with self-reported race and ethnicity labels as 4.5% Asian, 7.5% Black, and 88.0% White. For those with severe AS, TAVI was performed in 19.6% of Asian patients, 17.6% of Black patients and 24.9% of White patients; AVR was performed in 39.2% of Asian patients, 27.9% of Black patients and 32.8% of White patients. The mean time from severe AS diagnosis to TAVI was 0.69 years for Asian patients, 1.03 years for Black patients and 0.62 years for White patients (P=n.s.). The mean time to AVR was longer for Black patients (1.35 years) compared to Asian (0.49 years) and White patients (0.41 years, P<0.001). Survival in the overall cohort did not associate with ethnicity. However, in patients with severe AS, Black ethnicity was independently associated with increased mortality (hazard ratio=1.42, 95% CI=1.05-1.92, P=0.02).

**CONCLUSIONS AND RELEVANCE:** In patients with severe AS, Black patients experience lower rates of TAVI, longer time from diagnosis to AVR and higher rates of mortality, despite correction for socioeconomic deprivation. These data exhibit how AI technologies may be leveraged to shed light on health inequities, here showing that racial and ethnic disparities in AS persist in a universal healthcare system, and should stimulate strategies to address inequity.

**Key points:** *Question:* Do racial and ethnic disparities in the diagnosis, management, and outcome of aortic stenosis (AS) exist within a universal healthcare system?

*Finding:* In this retrospective cohort study using natural language processing enabled analysis of electronic healthcare record data of 5859 patients with aortic stenosis, we identified that in severe AS, Black patients experience lower rates of transcatheter aortic valve implantation (TAVI), longer times from diagnosis to surgical aortic valve replacement (AVR) and higher rates of mortality.

*Meaning:* Natural language processing may be used to identify health inequities. Here, we find racial and ethnic disparities in AS exist even in a universal healthcare setting.

## Introduction

Aortic stenosis (AS) is associated with considerable morbidity and mortality.[1, 2] Valve replacement, either by conventional surgery (AVR) or transcatheter aortic valve implantation (TAVI), is the treatment of choice in those with severe symptomatic AS.[3]

The interaction of race and ethnicity with inequality in care provision in AS is an area of emerging interest.[4] A limited body of evidence drawn from North American cohorts suggests the prevalence of AS to be lower in Black populations than White populations, while the risk factor profiles are paradoxically higher.[5, 6] In these cohorts, Black patients seem to be under-represented in those undergoing intervention for AS, possibly due to delays in diagnosis and treatment.[7, 8] For example, zip-code or county-level analyses in the USA found that rates of TAVI were lower in geographical areas with high proportions of Black patients with AS.[9]

Despite the different structures of healthcare provision in the USA and UK, health inequity exists in both countries, determined by the intersectionality of key factors including race and ethnicity (including structural racism), socioeconomic deprivation, and access to healthcare.[10–13] In addition to AS, health inequity has been noted to influence outcomes in a wide range of medical conditions.[14–17]

We hypothesised that race and ethnicity significantly impact AS diagnosis, management and outcomes within a universal healthcare system (**Figure 1a**). We conducted a retrospective study using longitudinal electronic health record (EHR) data from a tertiary UK hospital (**Figure 1b**). We used artificial intelligence (AI) methods – specifically natural language processing (NLP) - to identify all patients with a diagnosis of aortic stenosis. To evaluate the associations between race and ethnicity and clinical care for patients with AS we analysed the following outcome measures: (1) symptom and comorbidity burden at diagnosis, (2) time from symptom-to-diagnosis, and (3) the frequency, wait-time and mortality benefit of valvular interventions. Analyses were adjusted for patient age, sex, AS disease severity and we also explicitly controlled for the effects of socioeconomic deprivation.

**Figure 1.**
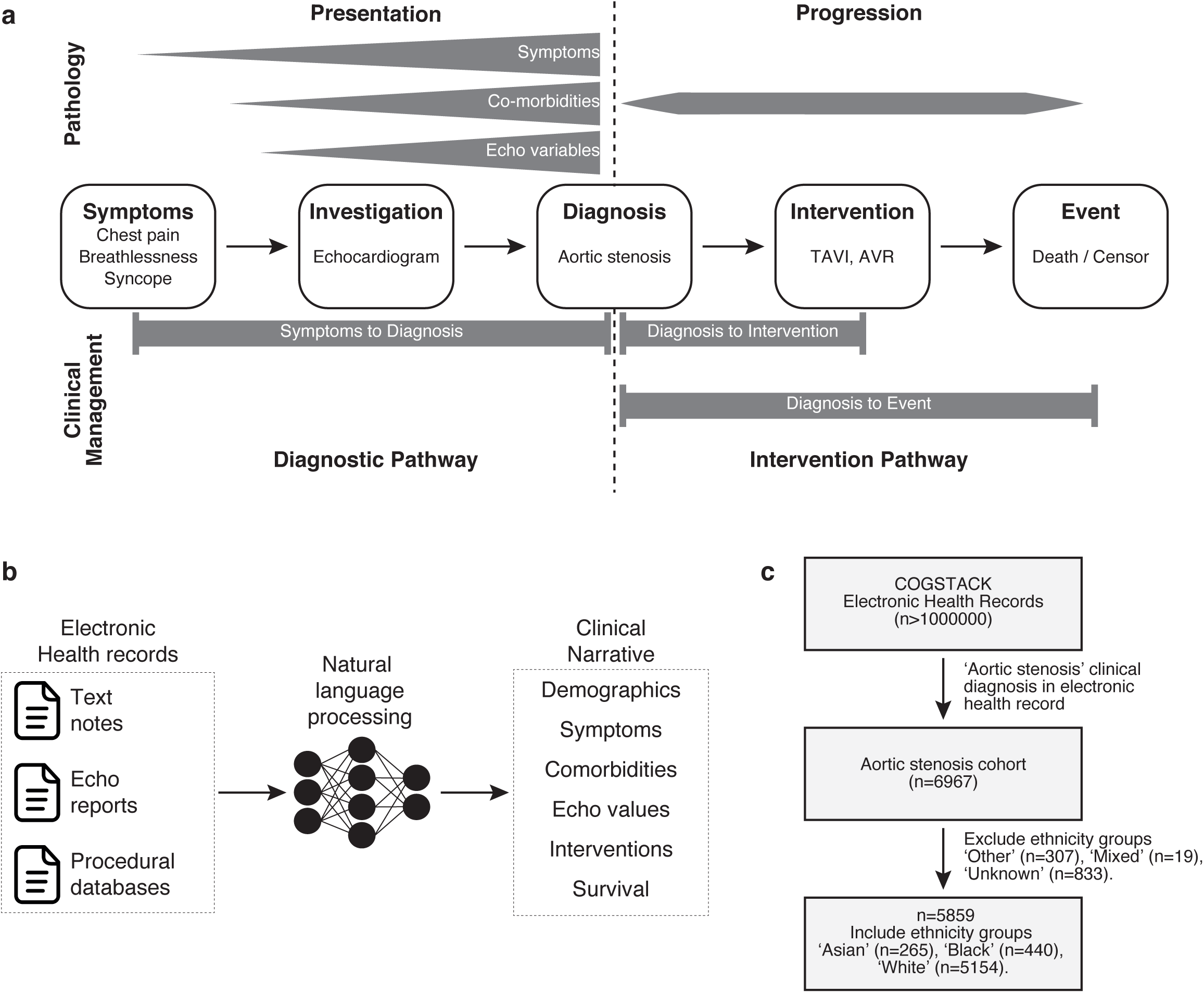
Cohort overview. a, Analysis framework to examine race and ethnicity differences in AS pathology and clinical pathways b, Natural language processing for extraction of clinical narratives from electronic health record c, CONSORT diagram of patient inclusion and exclusion criteria

## Methods

This project operated under London South-East Research Ethics Committee approval (18/LO/2048) granted to the King’s Electronic Records Research Interface (KERRI). This study complies with the Declaration of Helsinki.

### Study Cohort

EHR data were obtained from King’s College Hospital NHS Foundation Trust, a multi-site tertiary care hospital with a large urban hub and multiple suburban hospital sites in Southeast London. The clinical diagnosis of AS was identified using NLP (an AI technology that allows computers to interpret and understand human language) to extract the SNOMED-CT concept Aortic Valve Stenosis (concept ID: 60573004 with child concepts included) (**Supplementary Table S1**). The CogStack NLP pipeline allowed us to capture all mentions of the term ‘aortic valve stenosis’ in the electronic health records including the acronym ‘AS’ and synonyms such as ‘aortic stenosis’, while at the same time excluding cases of negation e.g. ‘this patient does not have aortic stenosis’. For a full description of the use of NLP in this study, please see **Supplementary Methods.** We included patients with an AS diagnosis between 1^st^ January 2010 to 31^st^ December 2020, with follow-up to 23^rd^ December 2022 which is used as the censorship date for survival analyses. We excluded patients under the age of 18 and those with self-ascribed ethnicity recorded as ‘mixed’, ‘other’ or ‘unknown’ (**Supplementary Table S2**).

### Data processing

Data (demographic, discharge summaries, clinical notes, transthoracic echocardiogram (TTE) reports) were retrieved and analysed in near real-time from the structured and unstructured components of the EHR using a variety of well-validated natural language processing (NLP) informatics tools belonging to the open-source CogStack ecosystem,[18] MedCAT,[19] and MedCATTrainer[20] deployed at King’s College Hospital NHS Foundation Trust. A detailed description is included in **Supplementary methods**.

### Study variables

The primary outcomes were symptom and comorbidity burden at AS diagnosis, time from first symptom to AS diagnosis, time from severe AS diagnosis to intervention, and all-cause mortality. Independent variables used in our analysis included race and ethnicity, age, sex, socioeconomic status, AS severity, symptoms, comorbidities, TTE variables, valvular interventions and survival outcomes. Categories for race and ethnicity were self-reported by patients and were extracted from the structured EHR along with age and sex. Socioeconomic status was established by mapping patient post code to The English Indices of Deprivation 2019 (https://www.gov.uk/government/statistics/english-indices-of-deprivation-2019), with an Indices of Multiple Deprivation (IMD) score calculated for each patient. Both structured and unstructured portions of the TTE reports were used to extract AS severity (mild, moderate and severe) of patients. Kaplan-Meier analysis was performed to confirm the expected stratification of patient survival based on mild, moderate and severe disease at one-year follow-up (**Supplementary Figure S1a**). Using MedCAT, SNOMED-CT concepts were extracted from clinical notes of AS patients, including cardiac symptoms, cardiovascular comorbidities, and non-cardiovascular comorbidities. Structured and unstructured portions of the TTE reports were also used to extract echocardiogram parameters including left ventricular ejection fraction, peak velocity, mean gradient, valve area and velocity ratio. The values from the TTE report with the closest date to the date of AS diagnosis were used to analyse echocardiogram differences at AS presentation. Valvular interventions were also extracted from the electronic health record, with manual validation demonstrating sensitivity to identify AVR as 95% and TAVI as 96% (**Supplementary Table S3**). To mitigate the confounding impact of the COVID-19 pandemic on intervention rates, TAVI and AVR data were accessed from a shortened time-frame (2010-2019). For a small number of patients receiving both AVR and TAVI procedures (n=12), the earlier procedure is used when stratifying for survival analyses. The date of diagnosis was defined as the first mention of AS in the electronic health record. The date of death was extracted from the electronic health record and this, or the last date of the data capture period, was used to calculate survival times.

### Statistical Analysis

Continuous variables were summarised as mean ± standard deviation (or standard error of the mean where appropriate) and categorical variables as counts with proportions. Analyses were adjusted as stated for patient age at diagnosis, sex, AS severity at diagnosis, and socioeconomic deprivation quintile (either ‘most deprived’ or ‘other’). Logistic regression was used to calculate odds ratios for being symptomatic or harbouring comorbidities at AS diagnosis. Kruskal-Wallis test was used to compare distributions of echocardiogram values. Linear regression was used to calculate model coefficients for time from symptom to AS diagnosis or time from severe AS diagnosis to intervention. Survival curves were plotted using Kaplan-Meier estimates, with log-rank tests used to assess significantly different survival times as stated. Cox regression was used to calculate hazard ratios for time from AS diagnosis to all-cause death or censoring. Schoenfeld residuals were plotted to evaluate the proportional hazard assumption. Statistical analyses were performed using R (version 4.3). All statistical tests were two-sided, unless otherwise stated, with P values <0.05 designated statistically significant.

## Results

### Demographics

From >1,000,000 individual patient records, we identified 6967 patients with a clinical diagnosis of AS (**Figure 1c**). 5859 patients with self-reported race and ethnicity coding data (**Supplementary Figure S1b**) with were included for analysis, composed of patients with an ethnicity label of White (n=5154), Black (n=440), or Asian (n=265).

### Baseline characteristics

Significant differences were observed in age, sex and socioeconomic status between race and ethnicity groups (**Table 1**). The average age at AS diagnosis for White patients (77.2y) and Asian patients (77.2y) was higher than for Black (74.8y). The majority of Black patients were female (60.7%), with the opposite seen in both Asian (37.7%) and White (45.3%). A higher proportion of Black patients (30.4%) were classified within the most deprived IMD quintile compared to White (11.8%) or Asian patients (10.5%, **Supplementary Figure S1c**).

**Table 1.**
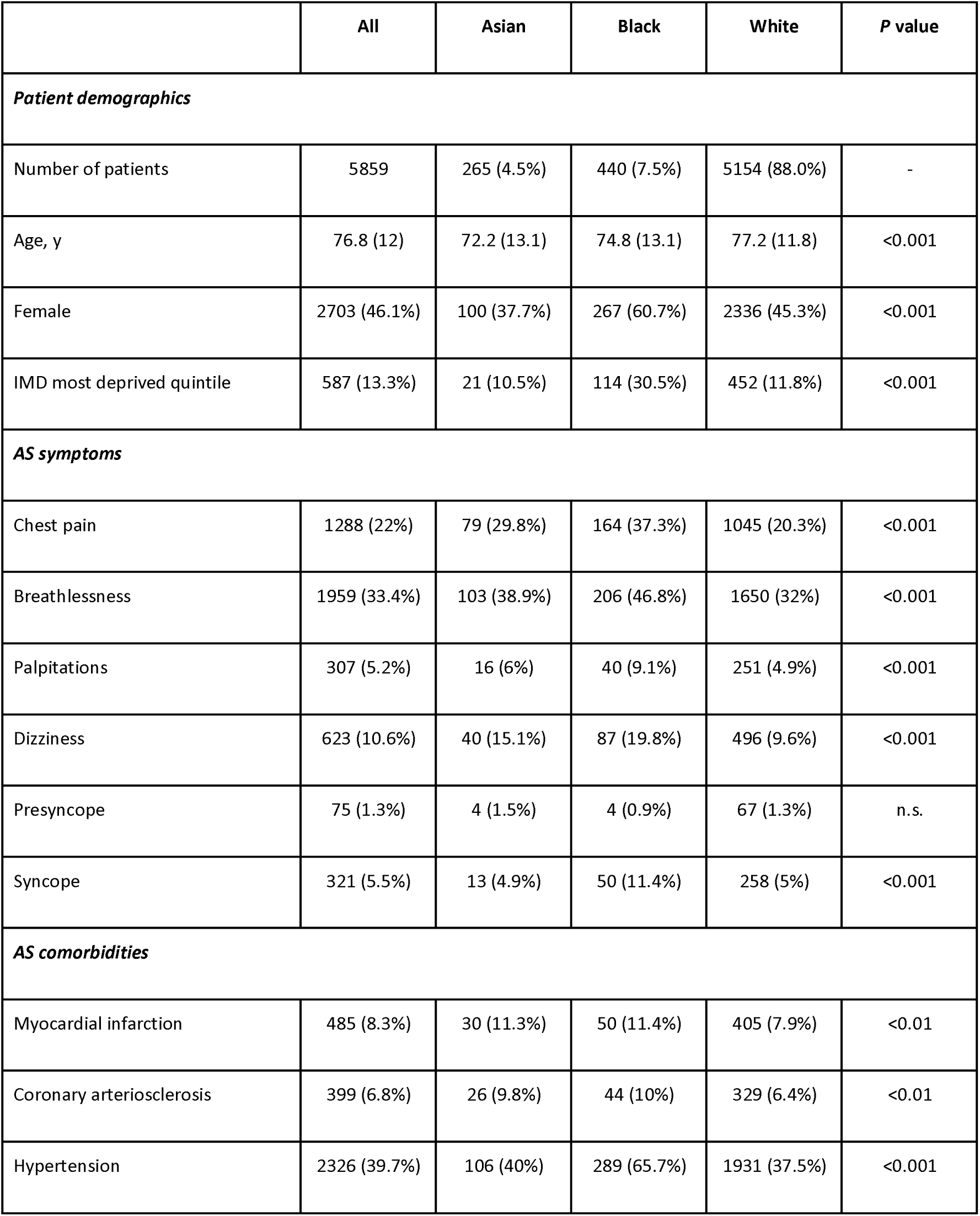
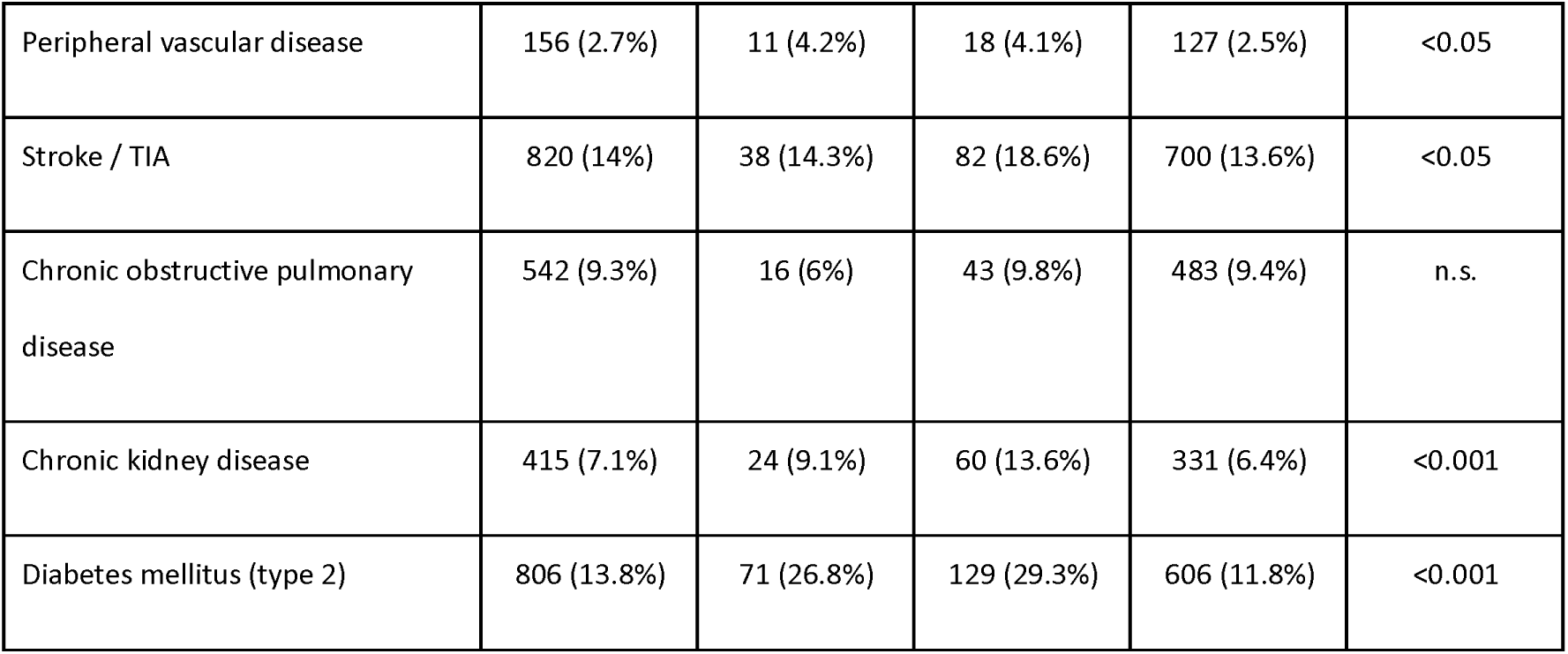
Baseline characteristics. Values are n (%) or mean ± standard deviation. Baseline characteristics were compared using a Pearson chi-square test for categorical variables and a Kruskal-Wallis test for continuous variables.

|  | All | Asian | Black | White | P value |
| --- | --- | --- | --- | --- | --- |
| ***Patient demographics*** |  |  |  |  |  |
| Number of patients | 5859 | 265 (4.5%) | 440 (7.5%) | 5154 (88.0%) | - |
| Age, y | 76.8 (12) | 72.2 (13.1) | 74.8 (13.1) | 77.2 (11.8) | <0.001 |
| Female | 2703 (46.1%) | 100 (37.7%) | 267 (60.7%) | 2336 (45.3%) | <0.001 |
| IMD most deprived quintile | 587 (13.3%) | 21 (10.5%) | 114 (30.5%) | 452 (11.8%) | <0.001 |
| ***AS symptoms*** |  |  |  |  |  |
| Chest pain | 1288 (22%) | 79 (29.8%) | 164 (37.3%) | 1045 (20.3%) | <0.001 |
| Breathlessness | 1959 (33.4%) | 103 (38.9%) | 206 (46.8%) | 1650 (32%) | <0.001 |
| Palpitations | 307 (5.2%) | 16 (6%) | 40 (9.1%) | 251 (4.9%) | <0.001 |
| Dizziness | 623 (10.6%) | 40 (15.1%) | 87 (19.8%) | 496 (9.6%) | <0.001 |
| Presyncope | 75 (1.3%) | 4 (1.5%) | 4 (0.9%) | 67 (1.3%) | n.s. |
| Syncope | 321 (5.5%) | 13 (4.9%) | 50 (11.4%) | 258 (5%) | <0.001 |
| ***AS comorbidities*** |  |  |  |  |  |
| Myocardial infarction | 485 (8.3%) | 30 (11.3%) | 50 (11.4%) | 405 (7.9%) | <0.01 |
| Coronary arteriosclerosis | 399 (6.8%) | 26 (9.8%) | 44 (10%) | 329 (6.4%) | <0.01 |
| Hypertension | 2326 (39.7%) | 106 (40%) | 289 (65.7%) | 1931 (37.5%) | <0.001 |
| Peripheral vascular disease | 156 (2.7%) | 11 (4.2%) | 18 (4.1%) | 127 (2.5%) | <0.05 |
| Stroke / TIA | 820 (14%) | 38 (14.3%) | 82 (18.6%) | 700 (13.6%) | <0.05 |
| Chronic obstructive pulmonary disease | 542 (9.3%) | 16 (6%) | 43 (9.8%) | 483 (9.4%) | n.s. |
| Chronic kidney disease | 415 (7.1%) | 24 (9.1%) | 60 (13.6%) | 331 (6.4%) | <0.001 |
| Diabetes mellitus (type 2) | 806 (13.8%) | 71 (26.8%) | 129 (29.3%) | 606 (11.8%) | <0.001 |

### AS presentation: symptoms and comorbidities

Breathlessness and chest pain were the most reported cardiac symptoms across all ethnicities (**Table 1**, **Supplementary Figure S2a**). A greater proportion of Black patients were symptomatic at diagnosis (62.0%) than White (42.1%) or Asian (45.7%) patients. Further, in a logistic regression model adjusted for age, sex, AS disease severity and socioeconomic deprivation, Black patients were significantly more likely to exhibit symptoms at AS diagnosis compared to White patients (adjusted odds ratio [aOR] = 1.92 [95% confidence interval = 1.48-2.51], P<0.001, **Supplementary Figure S2b-c**).

We observed significant racial and ethnic differences in a range of cardiometabolic comorbidities, most notably hypertension **(Table 1**, **Supplementary Figure S3a**). A higher proportion of Black patients (73.0%) were comorbid at AS diagnosis than White (47.8%) or Asian patients (49.8%, **Supplementary Figure S3b**). Black patients were significantly more likely to harbour comorbidities at AS diagnosis compared to White patients (aOR = 2.66 [95% CI=1.98-3.61], P<0.001, **Supplementary Figure S3c**) in a logistic regression model, adjusted for age, sex, AS disease severity and socioeconomic deprivation.

### Echocardiographic findings at AS diagnosis

At AS diagnosis, a higher proportion of White patients were diagnosed with severe disease (40.0%), compared with 24.3% of Black patients and 26.7% of Asian patients (**Supplementary Figure S4**). Examining echocardiographic parameters of left ventricular structure, Black patients displayed increased left ventricular thickness and higher left ventricular ejection fractions relative to White patients (**Supplementary Table S4**, **Supplementary Figure S4**).

### AS clinical management: diagnosis and intervention

As a proxy measure for diagnostic delay, we calculated the time interval between the first mention of a cardiac symptom and AS diagnosis. Analysing breathlessness, the mean symptom-to-diagnosis time for Black patients was 2.93 years (SEM=2.65-3.21), compared to 2.62 years (2.27-2.96) for Asian patients and 2.12 years (2.03-2.21) for White patients (**Figure 2a**). A linear regression model confirmed the longer symptom-to-diagnosis time for Black patients compared to White patients was robust to the effects of patient age, sex, AS disease severity and socioeconomic deprivation (coefficient=2.09 [95% CI=1.40-3.12], P<0.001, **Figure 2a, Supplementary Figure S5a**). For chest pain, the mean symptom-to-diagnosis time for Asian patients (mean=2.70 years, [SEM=2.37-3.03]) was longer than for Black (2.37 years, [2.14-2.60]) or White (1.69 years, [1.62-1.76]) patients (**Figure 2b**). This result was also significant in an adjusted linear regression model (coefficient=2.93 [95% CI=1.49-5.78], P<0.01, **Figure 2b, Supplementary Figure S5b**).

**Figure 2.**
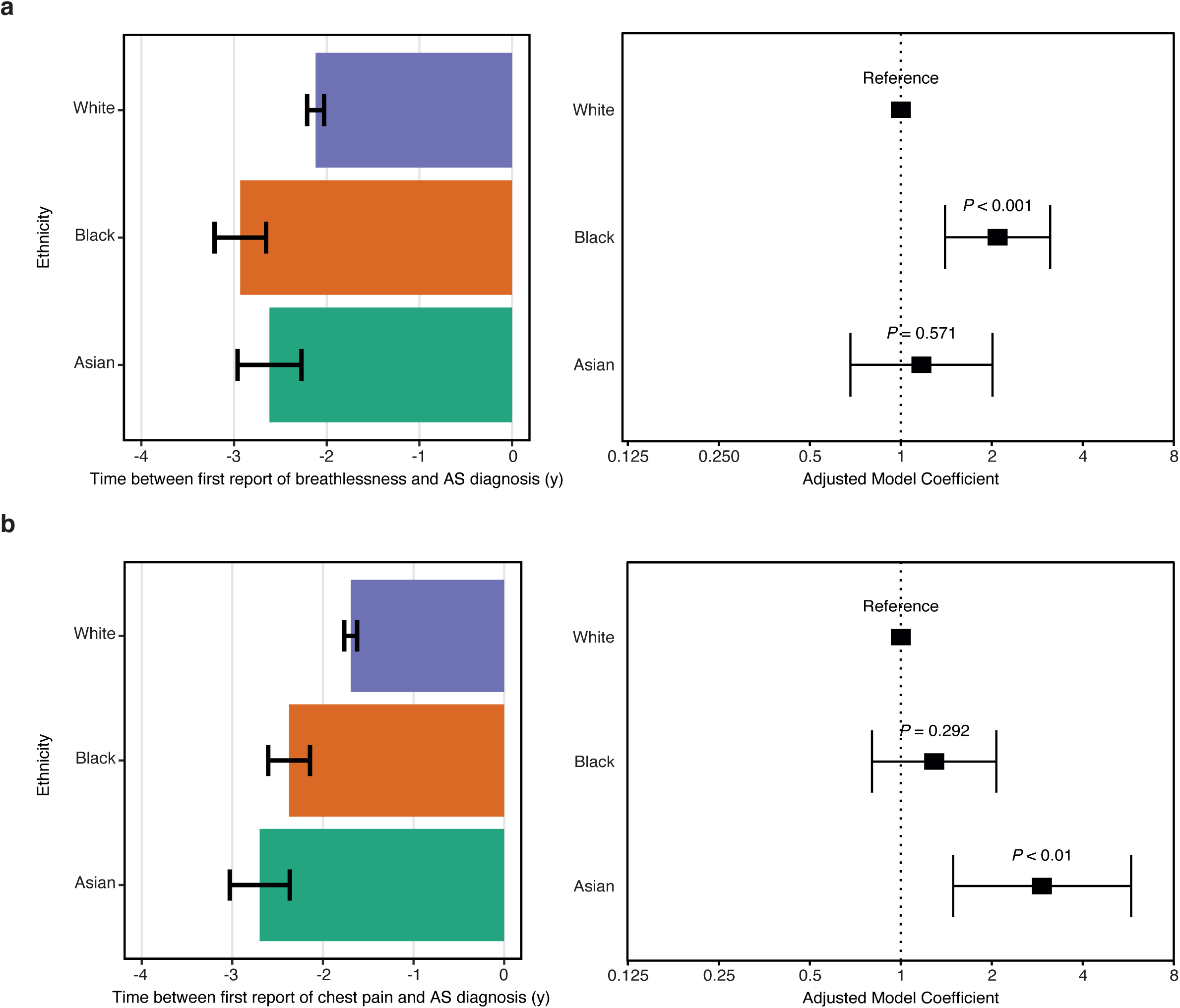
Lead-time from symptoms to AS diagnosis. a, Time between first report of breathlessness symptoms and AS diagnosis, stratified by ethnicity. b, Time between first report of chest pain symptoms and AS diagnosis, stratified by ethnicity. Bar plots (left) shows mean time difference. Error bars for bar plots represent the standard error of the mean. Forest plots (right) shows coefficients for a linear regression model adjusted for age, sex, AS disease severity and socioeconomic deprivation. Error bars for forest plots represent the limits of the 95% confidence interval for the model coefficient.

In a subgroup analysis of patients diagnosed with severe AS to identify race and ethnicity differences in the frequency of valvular interventions (TAVI or AVR), a higher proportion of White patients (24.9%) had a TAVI procedure than Black (17.6%) or Asian (19.6%) patients (**Figure 3a**). Similarly, higher proportions of White (32.8%) and Asian (39.2%) patients had a AVR procedure, with a lower proportion received by Black (27.9%) patients (**Figure 3a**). We also examined differences in intervention frequency according to socioeconomic status (**Figure 3a**).

**Figure 3.**
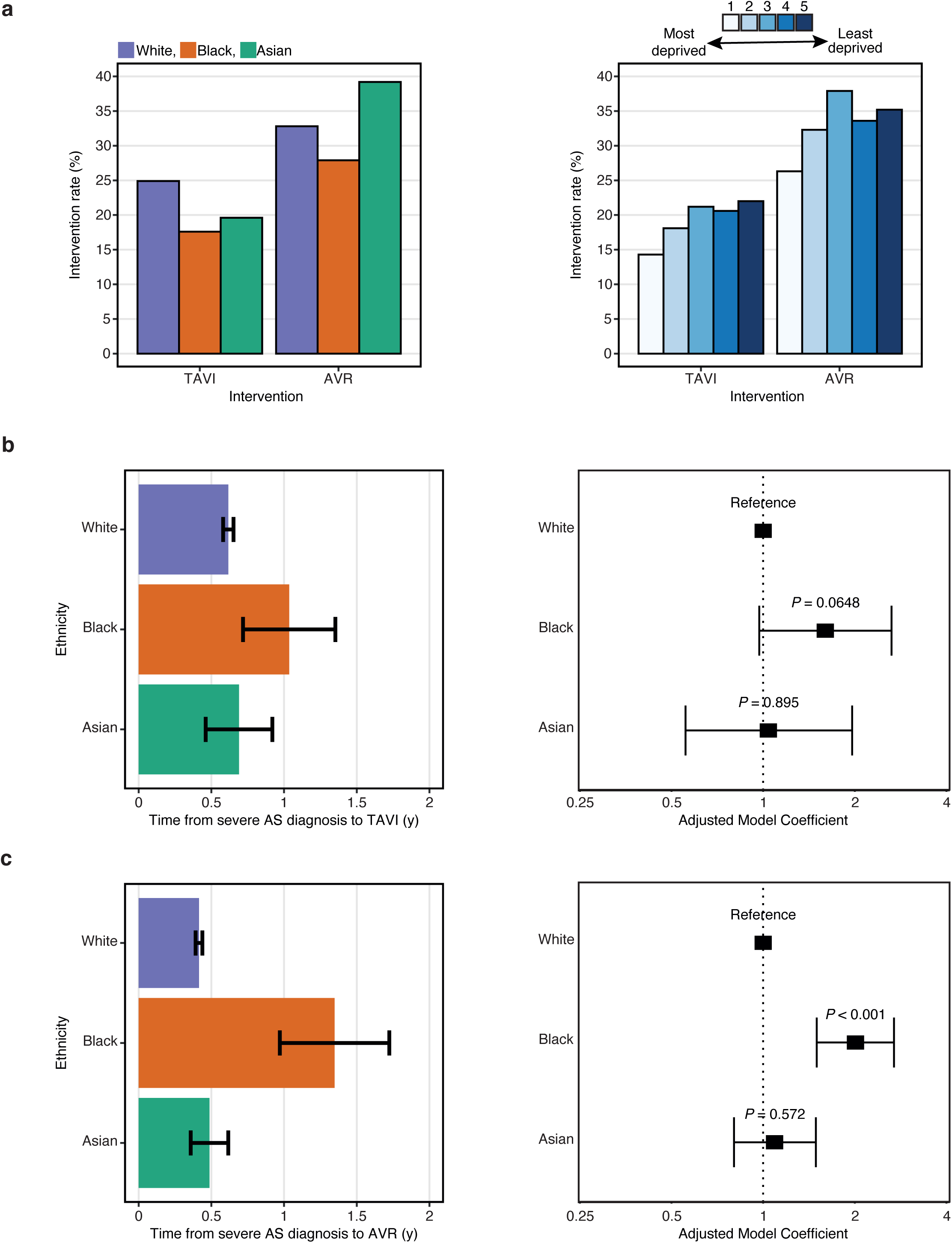
Intervention rate and wait-time. a, Bar plots showing the percentage of patients diagnosed with severe AS receiving an intervention (TAVI or AVR), stratified by ethnicity (left) or by deprivation quintile (right). b, Time between severe AS diagnosis and TAVI, stratified by ethnicity. Bar plot (left) shows mean time difference. Forest plot (right) shows coefficients for a linear regression model adjusted for age, sex and socioeconomic deprivation. c, Time between severe AS diagnosis and AVR, stratified by ethnicity. Bar plot (left) shows mean time difference. Forest plot (right) shows coefficients for a linear regression model adjusted for age, sex and socioeconomic deprivation. Error bars for bar plots represent the standard error of the mean. Error bars for forest plots represent the limits of the 95% confidence interval for the model coefficient.

We next calculated time intervals from the diagnosis of severe AS to valvular intervention in this patient subgroup. For TAVI procedures, the mean diagnosis-to-intervention time for White patients was 0.62 years (SEM=0.58-0.65), compared to 1.03 years (0.72-1.35) for Black patients and 0.69 years (0.46-0.92) for Asian patients (**Figure 3b**). In a linear regression model controlling for age, sex and socioeconomic deprivation, ethnicity was not significantly associated with time to TAVI (**Figure 3b, Supplementary Figure S6a**). For AVR procedures, the mean diagnosis-to-intervention time for Black patients (mean=1.35 years, [SEM=0.97-1.72]) was longer than for Asian (0.49 years, [0.36-0.62]) or White (0.41 years, [0.39-0.44]) patients (**Figure 3c**). The longer time to AVR in Black patients compared to White patients was statistically significant in a linear regression model adjusting for age, sex and socioeconomic deprivation (adjusted model coefficient=2.01 [95% CI=1.50-2.69], P<0.001, **Figure 3c, Supplementary Figure S6b**).

### Mortality

To explore the survival association of race and ethnicity in our cohort we examined five-year mortality post-diagnosis. This revealed no differences in survival times between patients of different ethnicities (**Figure 4a**). Multivariate Cox analysis confirmed no relationship between ethnicity and survival outcomes in the full cohort (**Figure 4b**), and also revealed that increasing age, male sex and socioeconomic deprivation were significantly associated with increased mortality (**Supplementary Figure S7**).

**Figure 4.**
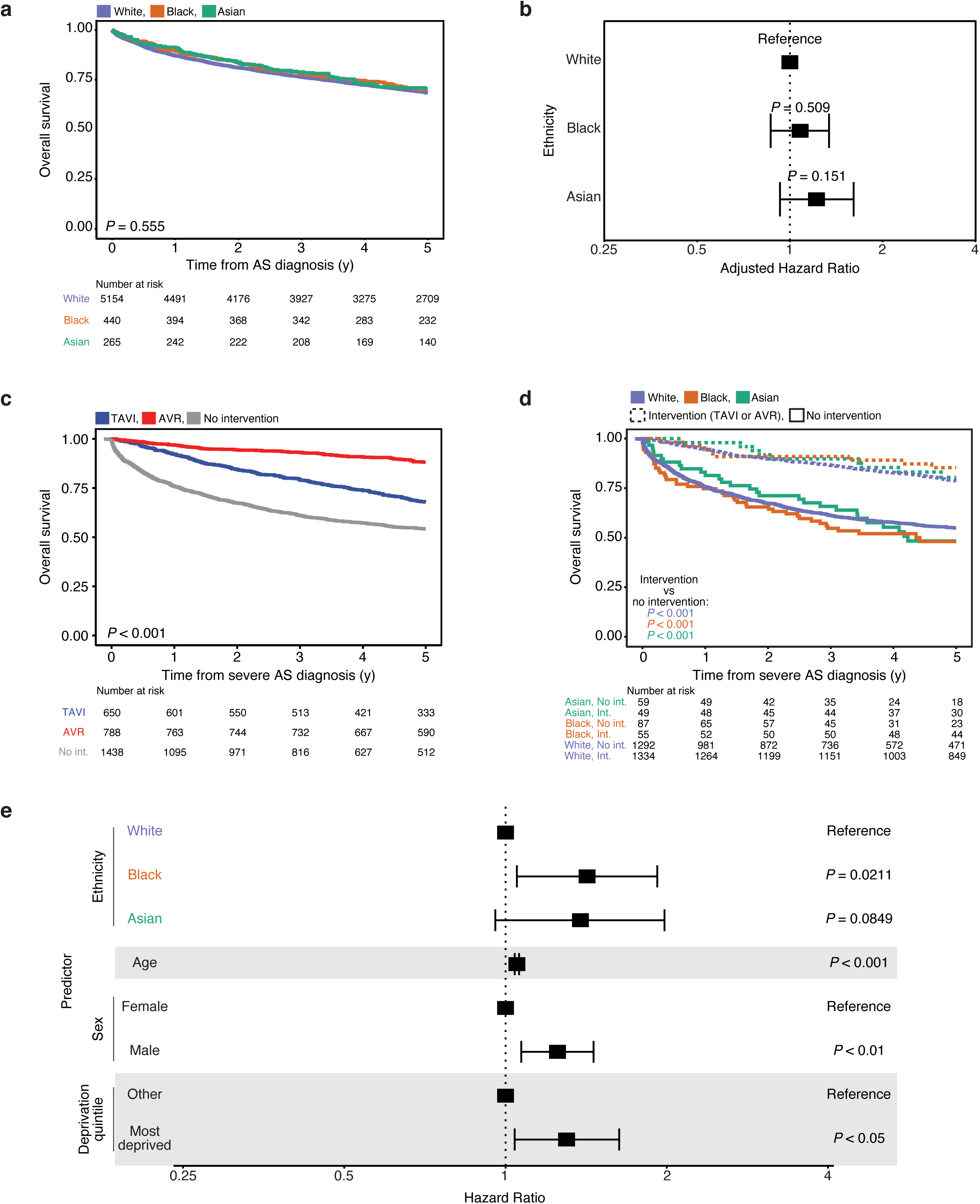
Mortality outcomes. a, Kaplan-Meier plot showing overall survival outcomes from AS diagnosis stratified by ethnicity. b, Forest plot showing adjusted hazard ratios for multivariate Cox analysis of overall survival outcomes from AS diagnosis stratified by ethnicity and adjusted for age, sex, socioeconomic deprivation and AS disease severity. c, Kaplan-Meier plot showing overall survival outcomes from severe AS diagnosis stratified by intervention. d, Kaplan-Meier plot showing overall survival outcomes from diagnosis of severe AS stratified by ethnicity and intervention status. e, Forest plot showing hazard ratios for multivariate Cox analysis of overall survival outcomes from diagnosis of severe AS stratified by ethnicity, age, sex and socioeconomic deprivation.

Next, we assessed survival times in patients diagnosed with severe AS, where valvular interventions may be indicated. Examining survival outcomes by intervention status, TAVI or AVR procedures were associated with a significant survival benefit (**Figure 4c**, P<0.001). We further evaluated this relationship sub-grouping patients by race and ethnicity, finding that any valvular intervention (either TAVI or AVR) was associated with a significant survival regardless of ethnicity (**Figure 4d**).

Lastly, we performed a multivariate Cox analysis of patients with severe AS. It was not possible to formally integrate intervention status (TAVI or AVR) in the Cox model as the proportional hazards assumption was violated (Schoenfeld residuals shown in **Supplementary Figure S8**). Black ethnicity was associated with increased mortality (**Figure 4e**, HR=1.42 [95% CI=1.05-1.92], P=0.0211). This analysis highlighted further patient features indicated with poorer survival outcomes (**Figure 4e)**, namely increased age (HR=1.05 [95% CI=1.04-1.06]), male sex (HR=1.25 [95% CI=1.07-1.46]), and socioeconomic deprivation (HR=1.30 [95% CI=1.04-1.63]).

## Discussion

This is the first study, to our knowledge, which uses natural language processing AI methods to augment quality measurement and improvement in Cardiology. Specifically, in this single-centre, retrospective, observational study we curated the EHR data of approximately 7000 patients with a clinical diagnosis of AS to investigate whether race and ethnicity are associated with differences in clinical presentation, management and outcomes. The UK setting of this study allowed us to evaluate whether racial and ethnic disparities persist in a universal healthcare system with explicit adjustment for the effects of socioeconomic deprivation. We find that Black and Asian patients are more likely to be symptomatic and harbour comorbidities at the point of AS diagnosis and wait longer from symptom onset to the clinical diagnosis of AS. Exploring valvular interventions in severe AS, Black patients have lower TAVI rates and wait longer for surgical AVR. Among patients with severe AS - and adjusting for age, sex and socioeconomic deprivation - Black patients experience significantly higher rates of mortality.

While we acknowledge that race and ethnicity are social constructs and there is no evidence that biological mechanisms in AS differ between ethnic groups,[24] the fact that Black ethnicity is associated with adverse prognosis in severe AS even after adjustment for socioeconomic deprivation, suggests the need for further exploration into the underlying mechanisms behind this finding, including considering factors such as structural racism. Strategies to actively address the treatment gap may include cultural training for healthcare providers and improved patient educational campaigns.[25]. As a research community there is a pressing need to increase the representation and reporting of race and ethnicity data in valvular heart disease clinical trials.[26]

We also find racial and ethnic variations in left ventricular (LV) echocardiographic parameters of patients with severe aortic stenosis. Compared to White patients, Black patients are more likely to have larger, thicker ventricles and higher LVEF values. These changes may reflect the higher rate of hypertension in Black patients relative to White patients, or ethnicity-specific LV remodelling patterns. Future research including time-series echocardiogram analysis could assess whether observed LV structural changes are static, reflecting a chronic hypertensive state, or are progressive due to LV remodelling with increasing AS severity.

### Strengths and limitations

The findings of this study are derived from a large, single-centre, retrospective database that uses AI-based methods to identify AS patients. The novel use of NLP to identify mentions of AS from the EHR is a strength of the study as previous approaches, relying on diagnostic coding or billing information, may under-code mild or early symptoms. Further, AI-based extraction of symptom, comorbidity and echo data directly from the clinical record facilitates high-resolution temporal analyses, including the calculation of symptom-to-diagnosis and diagnosis-to-intervention times. Conversely, a potential limitation of using NLP is that it may amplify any ethnicity-related biases in the clinical text recorded by clinicians.

We present the most comprehensive study examining the effects of race, ethnicity and socioeconomic deprivation in AS within a universal healthcare system. Our findings therefore add to the existing literature of North American studies, where potential payor biases may significantly influence patient and clinician behaviour.

The importance of integrating data on socioeconomic deprivation is indicated by several of our multivariate analyses, as IMD independently associates with being symptomatic and comorbid at diagnosis, and with mortality outcomes. However, there are limitations in the use of IMD as a metric of socioeconomic deprivation. Firstly, the choice of domains used to form the composite index is subjective and does not encompass the totality of socioeconomic deprivation. Secondly, it is important to recognise that IMD is a rank of an area’s deprivation compared to other areas within England (rather than the specific deprivation experienced by an individual within that area) and so represents only an indirect measure of the socioeconomic deprivation experienced at a patient-level.

This study has other important limitations. Firstly, our dataset is subject to the same potential biases as any other single-centre retrospective observational study. Our findings are observational in nature and relationships between variables do not imply causation. Due to the close geographic relation of London hospitals, it is possible that patients may have had prior AS investigation or management which is not reflected in the EHR accessed for this study. A further limitation is the use of high-level categories for race and ethnicity. Future research using larger datasets will allow for more granular categorisation (for example the separation of Asian patients into South Asian and East Asian categories) and the inclusion of categories not studied in this work (for example Middle Eastern or Hispanic patients) for added global relevance.

## Conclusion

In summary, we find racial, ethnic and socioeconomic differences in the presentation, diagnosis, and management of AS, in a universal healthcare system operating in the UK. In patients with severe AS, Black patients experienced higher rates of mortality. These data have important implications for the provision of equitable healthcare to the UK population.

## Data Availability

The datasets analysed during the current study are not publicly available due to hospital information governance regulations but are available from the corresponding author on reasonable request.

## List of non-standard abbreviations

AS: Aortic Stenosis
AVR: Surgical aortic valve replacement
IMD: Index of multiple deprivations
LVEF: Left ventricular ejection fraction
NLP: Natural language processing
TAVI: Transcatheter aortic valve implantation

## Access to data

KOG had full access to all data in the study and accepts responsibility for its integrity and the data analysis.

## Supplemental material contents

**Supplementary Table S1:** SNOMED concepts used to define the AS cohort

**Supplementary Table S2:** Supplementary data for patients with Mixed / Other / Unknown ethnicity.

**Supplementary Table S3:** Manual validation using independent procedural databases.

**Supplementary Figure 1:** Baseline characteristics

**Supplementary Figure S2:** Symptoms at AS diagnosis

**Supplementary Figure S3:** Comorbidities at AS diagnosis

**Supplementary Figure S4:** Echocardiogram variables stratified by ethnicity

**Supplementary Figure S5:** Lead-time from symptoms to AS diagnosis

**Supplementary Figure S6:** Intervention frequency and lead-time

**Supplementary Figure S7:** Mortality outcomes for all patients with AS

**Supplementary Figure S8:** Mortality outcomes for patients with severe AS

**Supplementary Methods: AS Cohort Identification using Natural Language Processing (NLP)**

