## SUPPLEMENTAL MATERIAL for "Natural Language Processing to Identify Racial and Ethnic Disparities in Aortic Stenosis"

**Racial and Ethnic Disparities in Aortic Stenosis within a Universal Healthcare System**

**Supplemental material contents:**

**Supplementary Table S1:** SNOMED concepts used to define the AS cohort

**Supplementary Table S2:** Supplementary data for patients with Mixed / Other / Unknown ethnicity**.**

**Supplementary Table S3:** Manual validation using independent procedural databases.

**Supplementary Table S4.** Echocardiogram values at time of diagnosis

**Supplementary Figure S1:** Baseline characteristics

**Supplementary Figure S2:** Symptoms at AS diagnosis

**Supplementary Figure S3:** Comorbidities at AS diagnosis

**Supplementary Figure S4:** Echocardiogram variables stratified by ethnicity

**Supplementary Figure S5:** Lead-time from symptoms to AS diagnosis

**Supplementary Figure S6:** Intervention frequency and lead-time

**Supplementary Figure S7:** Mortality outcomes for all patients with AS

**Supplementary Figure S8:** Mortality outcomes for patients with severe AS

**Supplementary Methods: AS Cohort Identification using Natural Language Processing (NLP)**

**Supplementary Table S1**

SNOMED concepts used to define the AS cohort

| **SNOMED ID** | **SNOMED Concept** |
| --- | --- |
| 60573004 | Aortic valve stenosis (disorder) |
| 19833008 | Nodular calcific aortic valve stenosis (disorder) |
| 703170006 | Prosthetic aortic valve stenosis (disorder) |
| 427515002 | Critical stenosis of aortic valve (disorder) |
| 194984004 | Aortic stenosis, non-rheumatic (disorder) |
| 194987006 | Aortic valve stenosis with insufficiency (disorder) |
| 703318003 | Neoaortic valve stenosis (disorder) |
| 81552002 | Rheumatic mitral valve insufficiency AND aortic valve stenosis (disorder) |
| 18546004 | Congenital stenosis of aortic valve (disorder) |
| 233863001 | Senile aortic stenosis (disorder) |
| 82355002 | Syphilitic aortic stenosis (disorder) |
| 72011007 | Rheumatic aortic stenosis (disorder) |
| 250973007 | Aortic stenosis with doming (disorder) |
| 44993000 | Rheumatic mitral valve and aortic valve stenosis (disorder) |
| 472847005 | Stenosis of fetal aortic valve (disorder) |
| 17759006 | Rheumatic aortic stenosis with regurgitation (disorder) |
| 703297006 | Prosthetic aortic valve stenosis and regurgitation (disorder) |
| 233862006 | Calcific aortic stenosis - bicuspid valve (disorder) |
| 703223000 | Postprocedural aortic valve stenosis (disorder) |
| 194735004 | Mitral insufficiency and aortic stenosis (disorder) |
| 703236000 | Non-rheumatic aortic valve stenosis with regurgitation (disorder) |
| 194733006 | Mitral and aortic stenosis (disorder) |
| 276790000 | Isolated aortic stenosis (disorder) |

**Supplementary Table S2**

Supplementary data for patients with Mixed / Other / Unknown ethnicity

Values are n (%) or mean (standard deviation).

|  | **All** | **Unknown** | **Other** | **Mixed** | ***P* value** |
| --- | --- | --- | --- | --- | --- |
| ***Patient demographics*** | | | | | |
| Number of patients | 1108 | 801 (72.3%) | 292 (26.4%) | 15 (1.4%) | - |
| Age, y | 75.2 (13.2) | 75.5 (13.6) | 74.5 (12) | 73.3 (11.5) | <0.05 |
| Female | 502 (45.3%) | 363 (45.3%) | 134 (45.9%) | 5 (33.3%) | n.s. |
| IMD most deprived quintile | 66 (11.8%) | 36 (9.6%) | 27 (15.4%) | 3 (27.3%) | <0.05 |
| ***AS symptoms*** | | | | | |
| Chest pain | 128 (11.6%) | 71 (8.9%) | 55 (18.8%) | 2 (13.3%) | <0.001 |
| Breathlessness | 205 (18.5%) | 129 (16.1%) | 69 (23.6%) | 7 (46.7%) | <0.001 |
| Palpitations | 35 (3.2%) | 18 (2.2%) | 15 (5.1%) | 2 (13.3%) | <0.01 |
| Dizziness | 57 (5.1%) | 35 (4.4%) | 19 (6.5%) | 3 (20%) | <0.05 |
| Presyncope | 6 (0.5%) | 5 (0.6%) | 0 (0%) | 1 (6.7%) | <0.01 |
| Syncope | 36 (3.2%) | 20 (2.5%) | 14 (4.8%) | 2 (13.3%) | <0.05 |
| ***AS comorbidities*** | | | | | |
| Myocardial infarction | 37 (3.3%) | 25 (3.1%) | 11 (3.8%) | 1 (6.7%) | n.s. |
| Coronary arteriosclerosis | 35 (3.2%) | 20 (2.5%) | 15 (5.1%) | 0 (0%) | n.s. |
| Hypertension | 267 (24.1%) | 172 (21.5%) | 87 (29.8%) | 8 (53.3%) | <0.001 |
| Peripheral vascular disease | 4 (0.4%) | 2 (0.2%) | 2 (0.7%) | 0 (0%) | n.s. |
| Stroke / TIA | 85 (7.7%) | 50 (6.2%) | 33 (11.3%) | 2 (13.3%) | <0.05 |
| Chronic obstructive pulmonary disease | 55 (5%) | 40 (5%) | 13 (4.5%) | 2 (13.3%) | n.s. |
| Chronic kidney disease | 25 (2.3%) | 19 (2.4%) | 6 (2.1%) | 0 (0%) | n.s. |
| Diabetes mellitus (type 2) | 78 (7%) | 51 (6.4%) | 25 (8.6%) | 2 (13.3%) | n.s. |

**Supplementary Table S3**

Manual validation using independent procedural databases.

| **Intervention** | **No. of patients receiving procedure** | | **Dataset coverage** |
| --- | --- | --- | --- |
|  | **Independent procedural database** | **Recovered by EHR search** |  |
| TAVI | 806 | 777 | 96% |
| AVR | 1230 (with AS)^*^ | 1170 | 95% |

In total there were 1834 AVR patients. The number of AVR patients with AS is an estimation after manual validation, by domain experts (KOG and colleagues), of a sample of 110 patients with AVR that were not covered in the EHR dataset presented. The result of the manual validation shows that 100 (91%) of the 110 patients did not have AS.

**Supplementary Table S4. Echocardiogram values at time of diagnosis**

Values are mean ± standard deviation.

|  | **All** | **Asian** | **Black** | **White** | ***P* value** |
| --- | --- | --- | --- | --- | --- |
| LVPWd (mm) | 1.18 ± 0.222 | 1.10 ± 0.2 | 1.25 ± 0.237 | 1.17 ± 0.219 | <0.001 |
| IVSd (mm) | 1.28 ± 0.246 | 1.19 ± 0.229 | 1.34 ± 0.271 | 1.28 ± 0.243 | <0.001 |
| LVIDd (cm) | 4.62 ± 0.775 | 4.48 ± 0.731 | 4.44 ± 0.76 | 4.65 ± 0.777 | <0.001 |
| LVIDs (cm) | 3.19 ± 0.856 | 3.09 ± 0.883 | 2.93 ± 0.805 | 3.22 ± 0.855 | <0.001 |
| LVEF (%) | 66.6 ± 15.3 | 68 ± 16.6 | 70.1 ± 14.1 | 66.1 ± 15.3 | <0.001 |
| LV mass (g) | 218 ± 72.1 | 187 ± 55.3 | 223 ± 82.3 | 219 ± 71.4 | <0.001 |
| LA volume (mL) | 74.7 ± 32.6 | 62.9 ± 33.1 | 72.0 ± 26.3 | 75.9 ± 33 | <0.001 |
| TR max velocity (m/s) | 266 ± 51 | 259 ± 47 | 269 ± 50.9 | 267 ± 51.2 | n.s. |

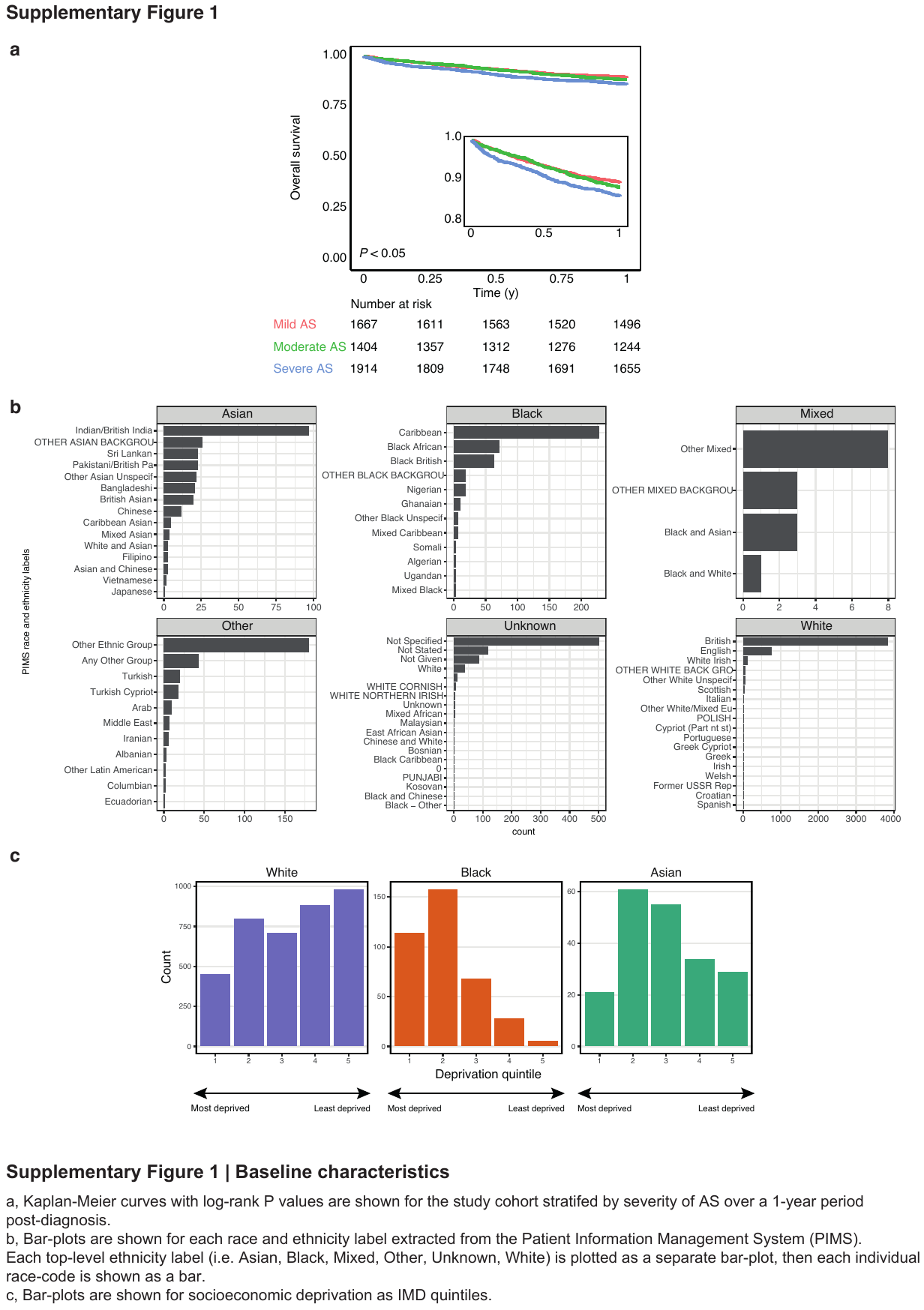

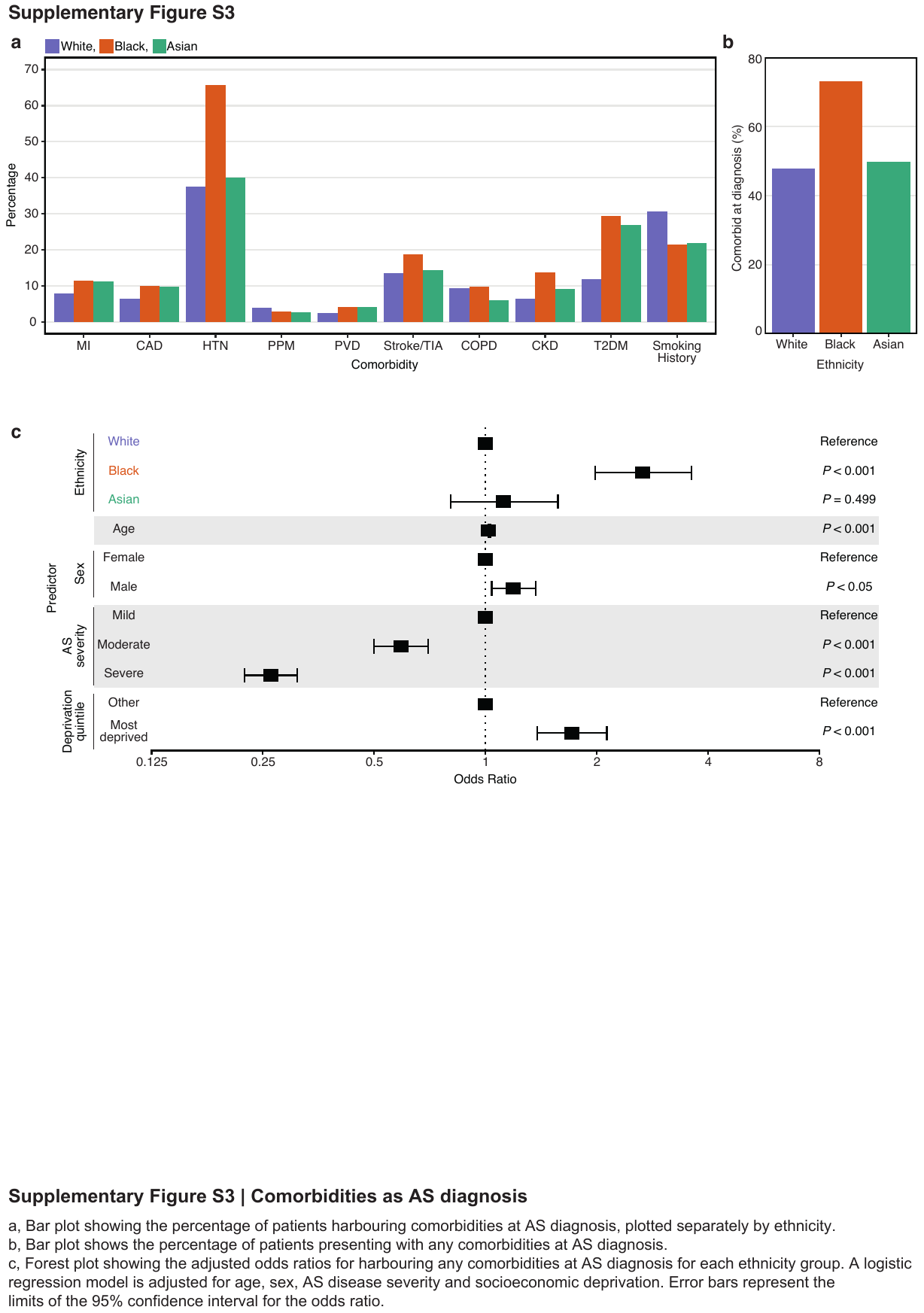

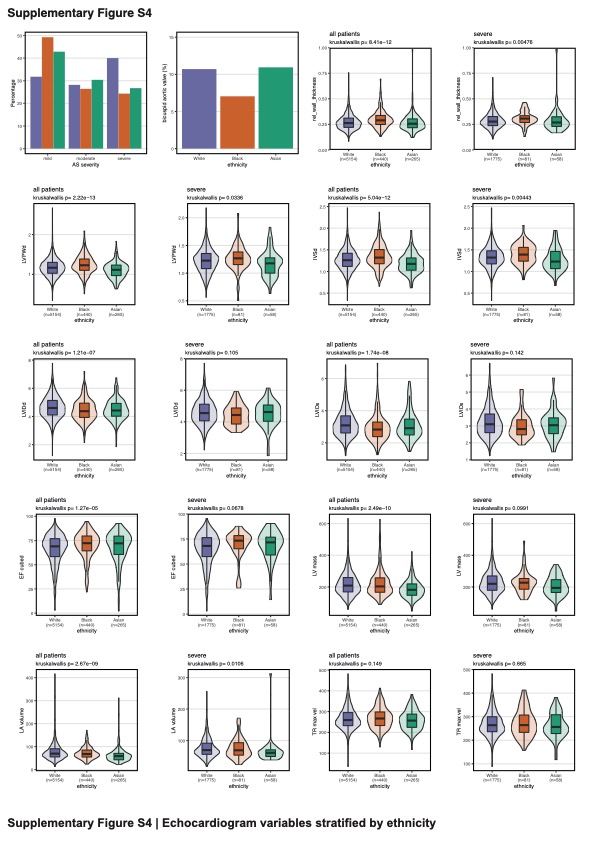

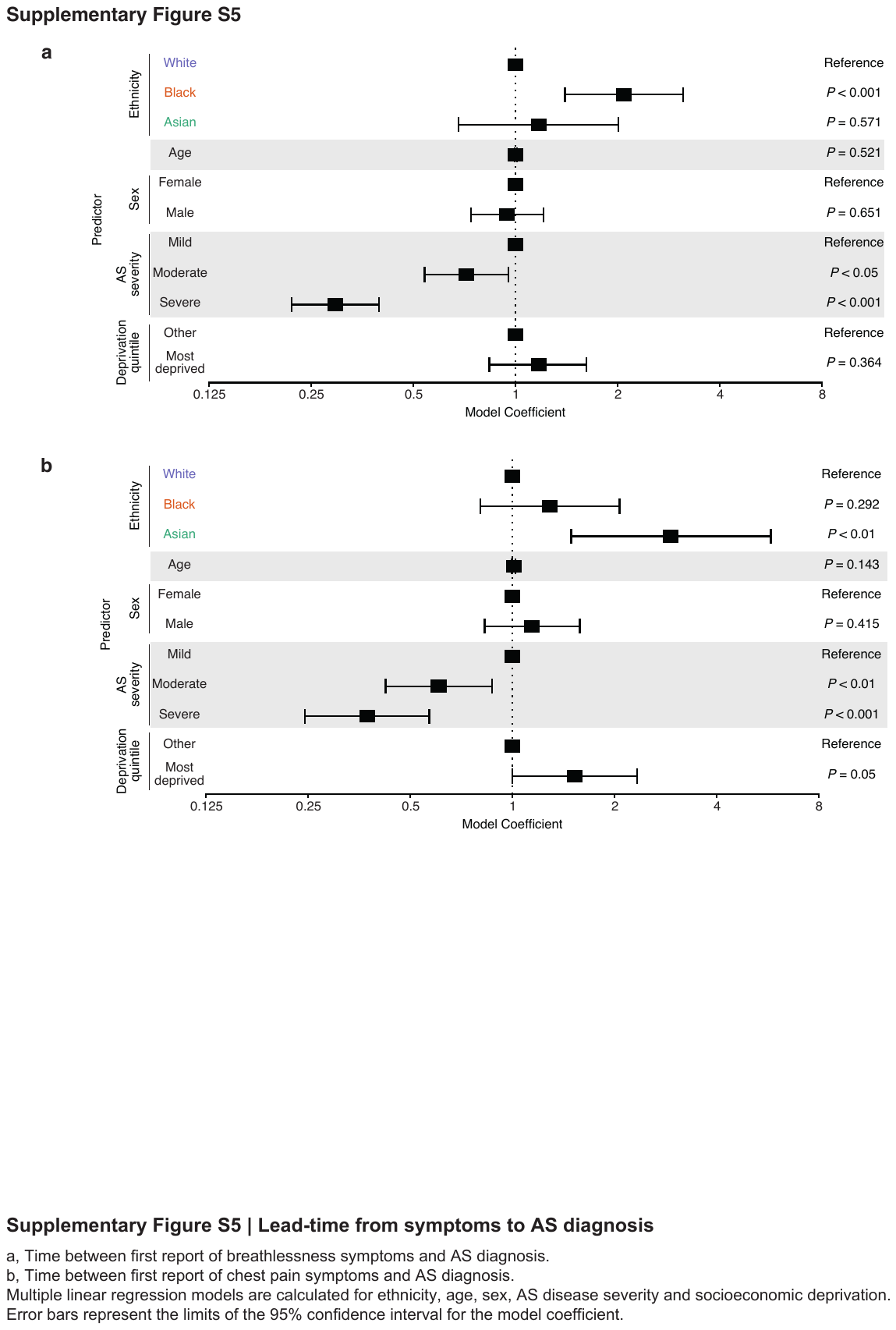

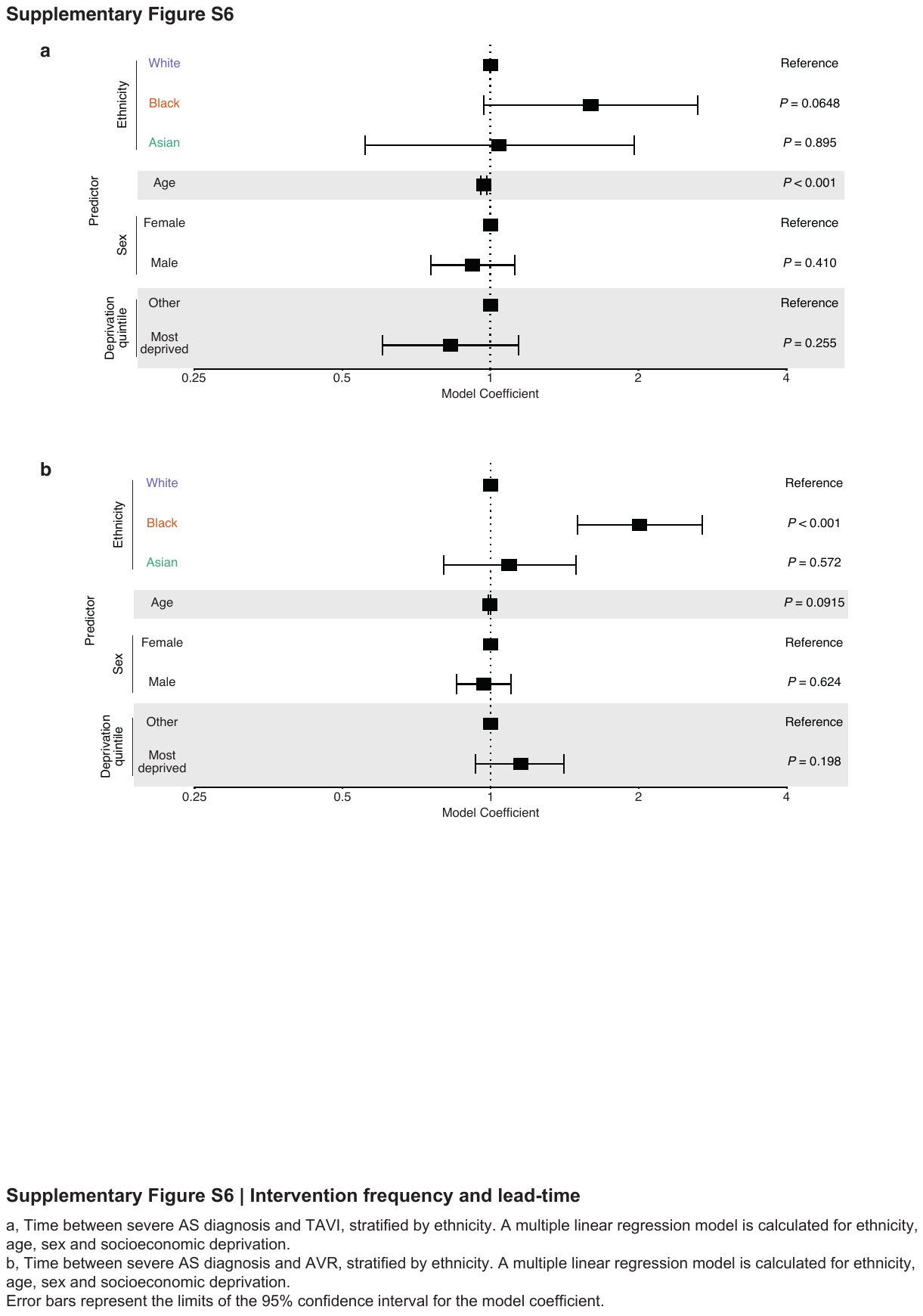

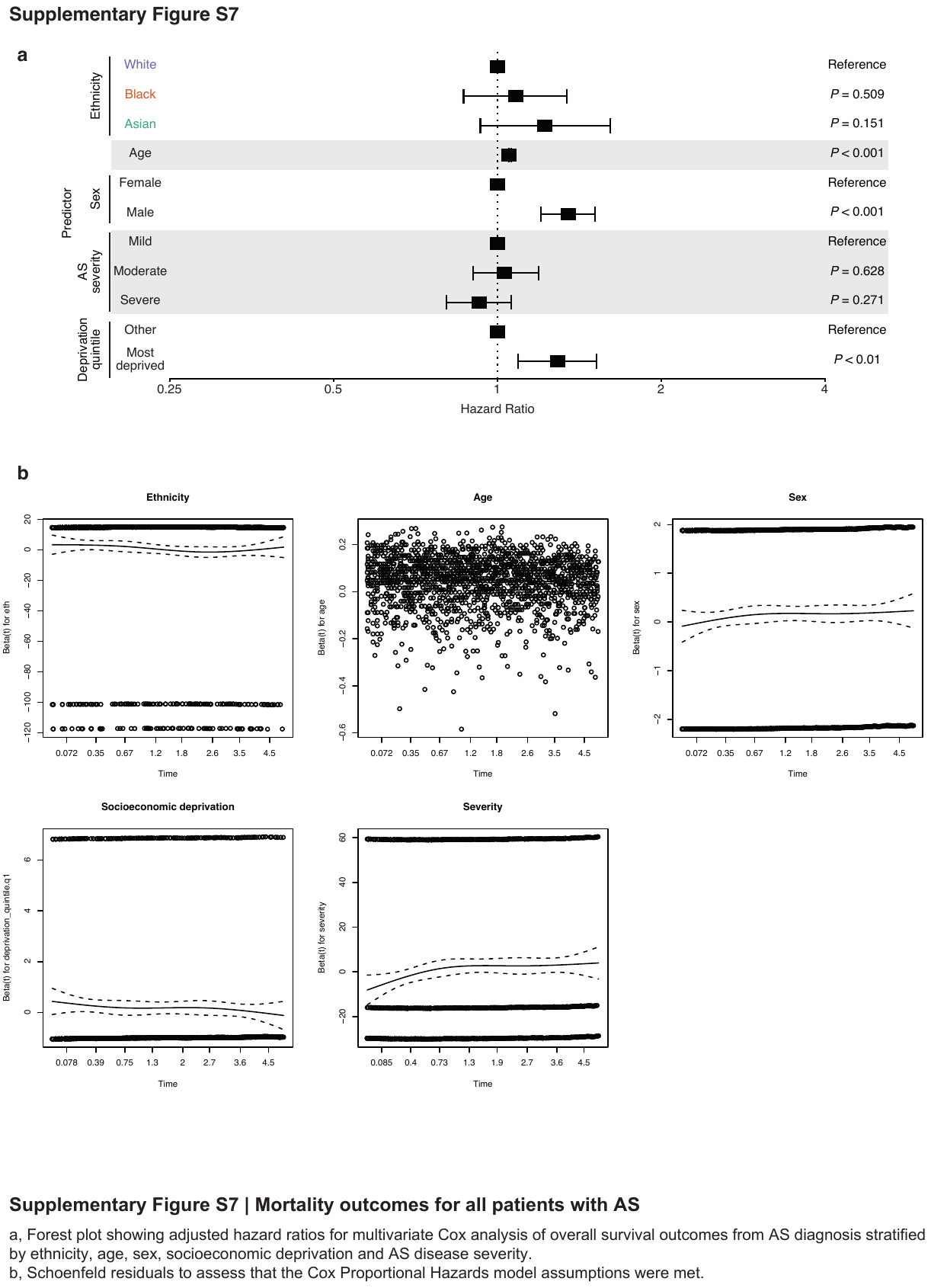

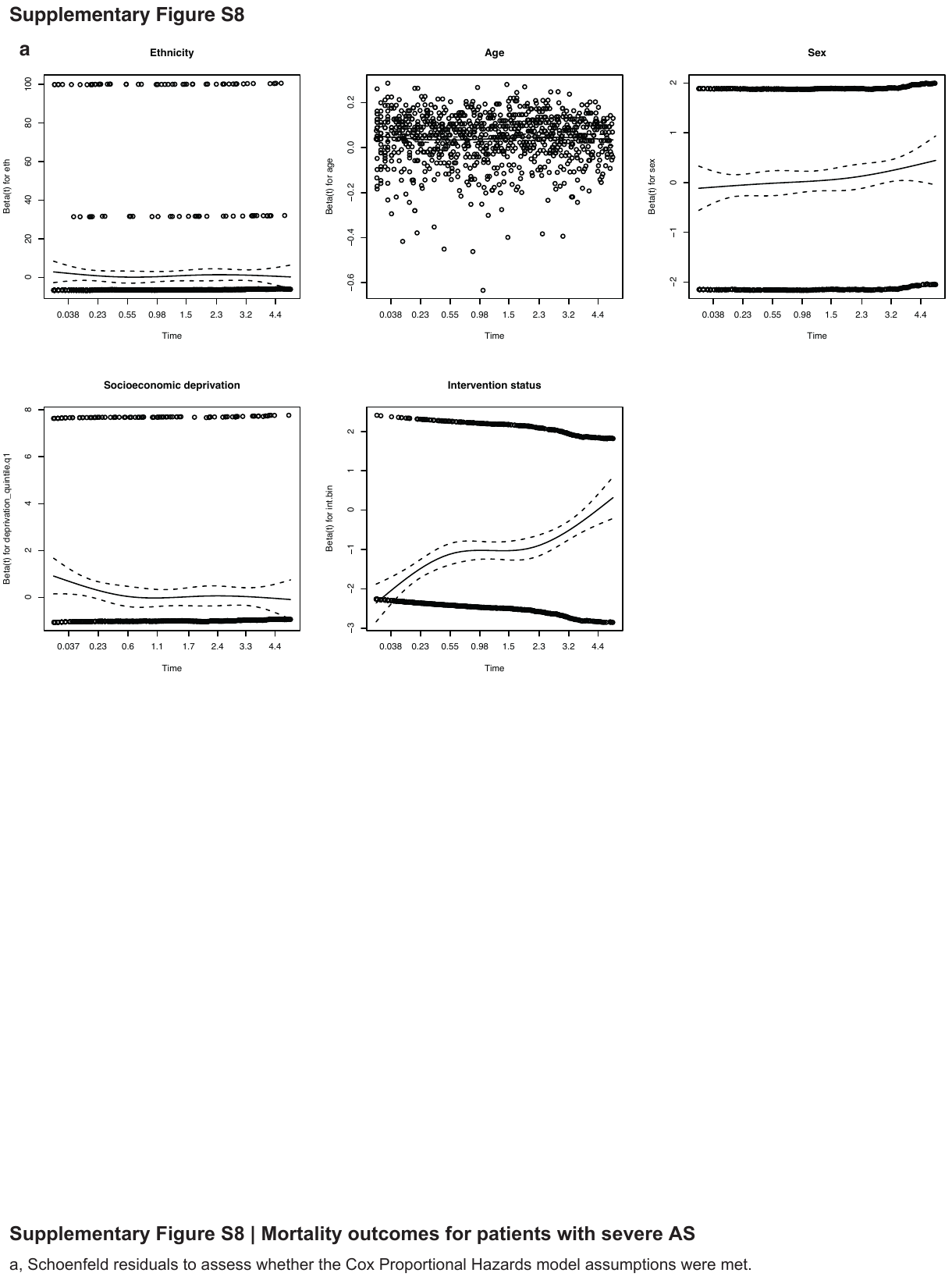

**Supplementary Methods: AS Cohort Identification using Natural Language Processing (NLP)**

The CogStack information retrieval system^1^ deployed at the King’s College Hospital NHS Foundation Trust collects structured and unstructured routinely collected data within the hospital. Structured data includes demographic (date of birth, sex, ethnicity, laboratory observations, etc) while unstructured data include free text documents of different types (clinical notes, discharge notifications, referral letters, etc). CogStack enables a single point of information retrieval related to patients’ electronic health records.

To facilitate effective and efficient search of clinical concepts in the unstructured data (free text documents), MedCAT is employed to annotate the text documents for all SNOMED concepts using machine learning approach. It is trained in both supervised and unsupervised fashion to achieve good accuracy for the detection of clinical concepts (F1 score > 0.9).^2^ MedCAT uses a novel concept disambiguation algorithm that learns clinical concept similarity via Word vector contexts and employs a Bidirectional Long-Short-Term-Memory (Bi-LSTM) model, a kind of deep learning models, for contextualisation (e.g., experiencer, negation, and temporal information). Correction of spelling mistakes is also performed during the annotation process.

MedCAT produced unsupervised annotations for all SNOMED‐CT concepts under parent terms Clinical Finding, Disorder, Organism, and Event with disambiguation, pre‐trained on MIMIC‐III.[21] Further supervised training improved detection of annotations and meta‐annotations such as experiencer (is the concept annotated experienced by the patient or other), negation (is the concept annotated negated or not) and temporality (is the concept annotated in the past or present) with MedCATTrainer. Meta‐annotations for hypothetical and experiencer were merged into irrelevant meaning that any concept annotated as either hypothetical or where the experiencer was not the patient was annotated as irrelevant. Performance of the MedCAT NLP pipeline for disorders mentioned in the text was evaluated on more than 5600 annotations for >265 documents by a domain expert and F1, precision and recall recorded.

**AS Cohort Identification**

We identified patients with a clinical diagnosis of AS using the SNOMED term “Aortic Valve Stenosis” (SNOMED ID: 60573004), with the following inclusion and exclusion criteria:

Inclusion criteria

- Patients were required to have at least two mentions of AS within the study timeframe i.e. 1 January 2010 to 31 December 2019.
- Age 18 years or over

Exclusion criteria:

- Patients with clinical notes containing only mentions of AS referring to individuals other than the patient i.e. within the family history
- Patients with clinical notes containing only negation mentions e.g. a statement in the clinical notes that the patient does not have aortic stenosis
- Patients with clinical notes containing only hypothesised references to AS.

**AS Severity Grading**

Severity grading of AS was extracted from both the echocardiogram reports and clinical notes. Within the echocardiogram reporting software at the King’s College Hospital NHS Foundation Trust, Xcelera, the written aspect of the report is not solely free text, but rather consisting of pre-coded sentences e.g. “No haemodynamically significant valvular aortic stenosis”. We used text matching to accurately extract AS severity from echocardiogram reports based on the presence of these pre-coded sentences.

In the clinical notes, where AS was mentioned (identified by MedCAT) we searched for the terms “mild” / “mild to moderate” / “moderate” / “moderate to severe” / “severe” immediately before the AS mentions. To avoid ambiguity, we grouped together “mild” and “mild to moderate” AS as a single mild AS group, with “moderate” and “moderate to severe” AS as a single moderate AS group.

Where data on AS severity was available in the echocardiogram report, this was used at the primary source. If no data was available in the echocardiogram report, data from the clinical notes was used.

1.	Jackson R, Kartoglu I, Stringer C, Gorrell G, Roberts A, Song X, Wu H, Agrawal A, Lui K, Groza T, et al. CogStack - experiences of deploying integrated information retrieval and extraction services in a large National Health Service Foundation Trust hospital. *BMC Med Inform Decis Mak*. 2018;18:47. doi: 10.1186/s12911-018-0623-9

2.	Kraljevic Z ST, Shek A, Roguski L, Noor K, Bean D, Mascio A, Zhu L, Folarin AA, Roberts A, Bendayan R, Richardson MP, Stewart R, Shah AD, Wong WK, Ibrahim Z, Teo JT, Dobson RJB. Multi-domain Clinical Natural Language Processing with MedCAT: the Medical Concept Annotation Toolkit. *Artif Intell Med*. 2021;117:102083.
